# Time-to-sputum culture conversion, treatment outcomes, and associated factors among multidrug-resistant tuberculosis patients in the Sidama region, Ethiopia: A retrospective follow-up study

**DOI:** 10.1101/2025.06.21.25330054

**Authors:** Wolde Abreham Geda, Kebede Tefera Betru, Tarekegn Solomon, Solomon Daniel

## Abstract

**Background:** Sputum-culture conversion is critical to follow-up treatment effectiveness in multidrug-resistant tuberculosis patients. However, the evidence regarding time-to-culture conversion, treatment outcomes, and the associated factors was sparse and inconsistent.

**Objective:** This study aimed to determine the time-to-culture conversion, treatment outcomes, and associated factors among multidrug-resistant tuberculosis in the Sidama Region from April 1 to December 31, 2024.

**Methods:** We conducted a retrospective follow-up study on 346 patients who enrolled from January 2013 to June 2024. The researchers collected data from the patient’s medical records using the standardized forms, entered the data, and analyzed it using SPSS 26 software. The researchers performed the analysis using the Kaplan-Meier model time-to-culture conversion, Cox proportional hazard ratio, and logistic regression models to assess factors associated with time-to-culture conversion and treatment outcomes, respectively. We considered an adjusted hazard ratio with a 95% CI and a P-value < 0.05 to decide the significance.

**Results:** Among the participants, 302 (87.3%) achieved culture conversion in a median time of 76 days (95% CI: 71-79, p < 0.05); 36 patients (10.4%) displayed a delayed conversion, with a median conversion time of 141 days while, 8 (2.3%) patients did not show culture conversion. Among the participants, 32 patients (9.2%) were not evaluated for their treatment outcomes. The treatment success rate was 234 (67.6%). The rates of cure, treatment completion, death, treatment failure, and loss to follow-up were 177 (51.2%), 57 (16.5%), 23 (6.6%), 5 (1.4%), and 52 (15%), respectively. Patients with a history of previous treatment failure had worse treatment outcome with an aHR of 6 (1-27), 95% CI, P = 0.002.

**Conclusions:** Most of the patients (87.3%) achieved culture conversion within three months. However, patients with a history of retreatment faced worse outcome, pointing to serious treatment follow-up to the patients.

## Introduction

Tuberculosis (TB) is the deadliest infectious disease, but it is curable if the patients get the appropriate treatment regimen. Roughly TB infected a quarter of the global population. However, most infected will not develop active TB disease, and some individuals may recover due to their immune response. TB has become a top killer among infectious diseases.^1^

Globally, approximately 10.6 million people were diagnosed with tuberculosis in 2022.2 Among people who were diagnosed with TB, 400,000 people additionally acquired Multidrug-resistant/ Rifampicin mono-resistant-TB (MDR/RR-TB). Ethiopia is one of the 30 countries with a high burden of tuberculosis (TB) and TB-HIV co-infection. In 2022, the estimated annual incidence of TB was 126 cases per 100,000 people, accompanied by a death rate of 17 per 100,000. Although Ethiopia has transitioned out of the list of countries with high multidrug-resistant tuberculosis (MDR-TB) burden since 2020, drug-resistant TB continues to pose a significant public health challenge. In 2022 alone, there were an estimated 2,000 new cases of RR/MDR-TB, with a considerable number of cases remaining undetected.^1^

The rise of drug-resistant TB (DR-TB) has intensified this public health issue and remains a persistent threat. The resistance to rifampicin, which is the most effective first-line treatment, has posed a great concern and when resistance occurs to both rifampicin and isoniazid, the condition is classified as multidrug-resistant TB (MDR-TB). Both MDR-TB and rifampicin-resistant TB (RR-TB) require treatment with second-line medications.^3^

Over the years, global and local stakeholders have made vast efforts to prevent and control transmission, reduce mortality, and mitigate the associated disease burden. The global campaign undertaken to combat tuberculosis includes directly observed short-course treatment, the Stop TB Strategy, the current End-TB strategy, etc. Consequently, there were encouraging signs of progress. However, TB remains one of the public health challenges, especially in developing countries.^2^

Proper diagnosis and treatment follow-up is needed to ensure better treatment success rate and halt the emergency of MDR-TB or extensively drug resistant (XDR) TB.^4^

MDR-TB treatments are weak in clearing the bacteria but cause toxic side effects to the patients who receive the drugs. On the contrary, the treatment success rate would be lower if the treatment regimen is not accompanied by cautious patient adherence, periodic treatment monitoring, using an appropriate tool, time to sputum culture conversion.^4^

Time-to-sputum culture conversion is a surrogate indicator of the treatment effectiveness and final treatment outcomes. It is a tool to determine the duration of treatment for MDR-TB patients. During the intensive phase, the MDR-TB treatment continued for at least four months after the culture conversion or eight months, whenever longer.^5^

MDR-TB treatment takes longer, 18-20 months, and it is very costly. Consequently, the treatment process consumes the resources, energy, and time of a patient and health worker. The periodic culture evaluation is a milestone towards the end of the treatment outcomes. In turn, the final evaluation of the treatment outcomes highlights the quality of service rendered to the patients.^6^

Moreover, understanding factors associated with time-to-sputum culture conversion and associated factors related to the treatment outcomes helps to improve patient outcomes by altering their impact.^7^

Therefore, predicting the initial-time to culture conversion is crucial in planning and implementing respiratory isolation, to determine the duration of treatment, regimen modification and ascertaining the overall length of MDR-TB treatment.^6^

This study generated additional and updated evidence about when the sputum culture would convert from positive to negative after the initiation of MDR-TB treatments and the associated factors for time-to-sputum culture conversion.

As the treatments for multidrug-resistant tuberculosis is changing overtime, this study assessed for a potential change in treatment outcomes among the patients receiving different treatment regimens, and the associated factors.

Consequently, the current study would also generate up-to-date evidence for clinicians and policymakers to understand the time length in which the MDR-TB patient would clear from mycobacteria. The evidence generated would also assist clinicians in determining the duration of anti-TB treatment for the patient. Understanding time-to-sputum culture conversion among MDR-TB patients would also help to know how long a patient would remain infectious after initiation of the second-line anti-TB treatment. As a result, this assists in cutting the spread of the disease, especially among households and close contacts.

## Methods

### Study area

The study was conducted in the Sidama region of Ethiopia, situated in the central-southern part of the country. This region shares borders with two other significant regions: the Oromia region to the north, northeast, south, and southeast, and the South-Ethiopia region, which lies to the south.

Geographically, the region is positioned between 6° 14’ and 7° 15’ North Latitude and 37° 9’ to 39014 East Longitude. The total area of the region encompasses approximately 6,690.51 square kilometers, of which 97.71% is designated as land and 2.29% consists of water bodies. The region has an extensive boundary length of 6,008 kilometers, with the most significant portion (86%) bordering Oromia and the remaining 14% adjacent to the south Ethiopia Region. The shape of Sidama is notably compact, extending predominantly in the southeast direction.

According to the Central Statistical Agency (CSA) report based on the 2007 census, the population of the Sidama region was recorded at 4,647,672 individuals. The region is equipped with a healthcare infrastructure that includes 21 hospitals, 138 health centers, and 553 health posts. Notably, among the 21 hospitals, only one—Yirgalem Hospital—provided treatment for multidrug-resistant tuberculosis (MDR-TB). As a result, all MDR-TB patients in the region are referred to Yirgalem Hospital for the initiation of treatment, while the other hospitals and health centers offer follow-up care services to those patients.

### Study design and period

We carried out a retrospective follow-up study on tuberculosis patients with rifampicin mono-resistant or MDR-TB and sputum culture-positive patients. The patients were registered from January 1, 2013, to June 30, 2024, from the Sidama region of Ethiopia at the Yirgalem treatment initiation center. The researchers accessed the patient data anonymously, using their medical and multidrug-resistant TB record number, from April 1 to December 31, 2024.

### Source and study population

The source population was the MDR/RR-TB patients in the Sidama region.

The study population consisted of all culture-positive rifampicin-resistant or MDR-TB patients who started anti-TB treatment.

### Inclusion and exclusion criteria

MDR/RR-TB patients aged over fifteen years who were culture positive at the start of treatment were included in the study.

The exclusion criteria for the study were patients aged less than fifteen years, patients with incomplete data, patients with culture-negative results at baseline, and patients diagnosed with MDR/RR-TB who did not begin anti-TB treatment.

### Sample size decision

The sample size for this study was decided using OpenEpi Version 3 software, which is specifically designed for cohort studies.^8, 9^

The parameters used in the calculation are provided below (Table 1).

**Table 1:** The Sample size determination for time-to-sputum culture conversion, treatment outcomes, and associated factors among multidrug-resistant tuberculosis patients in the Sidama region (n= 346).

| Parameters | Level |
| --- | --- |
| Two-sided significance level(1-alpha): | 95 |
| Power(1-beta, % chance of detecting): | 80 |
| Ratio of sample size, Unexposed/Exposed: | 1 |
| Percent of Unexposed with Outcome: | 5 |
| Percent of Exposed with Outcome: | 7.2<sup>10</sup> |
| Odds Ratio: | 1.5 |
| Risk/Prevalence Ratio: | 1.4 |
| Risk/Prevalence difference: | 2.2 |

The largest sample size for a single-arm cohort study was = 1977, but the total eligible RR/MDR-TB population in the region during the study period was 420. Therefore, we considered the entire RR/MDR-TB population for the study. However, 74 patients were removed for their exclusion criteria, leaving the final sample size to 346.

### Data collection and sampling technique

After determining the sample size, the sampling frame was created, and a systematic random sampling technique was utilized to collect the sample. Data were then gathered from the selected participants using a standardized data collection form, which ensured consistency and reliability in the data acquisition process.

### Data Integrity

Before the comprehensive implementation of data collection, 5% of the patient medical records in the study area were reviewed. Insights gained from this initial assessment led to revisions of the data collection tool before the actual data gathering commenced. Data collectors and supervisors were chosen from among health professionals with clinical expertise, and they underwent two days of training on the data extraction tool. To ensure data completeness, a meticulous review was conducted daily before personnel left the facility. Data cleaning was executed by conducting frequency analyses in SPSS 26 to verify the plausibility of each variable, identify any missing items, and assess their acceptability. Furthermore, random checks were performed on 5% of the cases to maintain data integrity.

### Dependent and independent variables

The dependent variables time-to-sputum culture conversion, treatment outcomes consisting of cure, completion, death, loss to follow-up, treatment failure, not evaluated and associated factors with time-to-culture conversion and treatment outcomes included different types of drug resistance: rifampicin mono-resistance, isoniazid mono-resistance, and multidrug resistance. Additionally, various treatment regimens were considered, such as short-term treatment, long-term treatment, standardized treatment, individualized treatment, and the Bedaquiline, Pretomanid, Linezolid, and Moxifloxacin (BPaLM) regimen.

Demographic variables such as sex, age, and residence, along with clinical variables like the patient registration group (RR, MDR, or Isoniazid (INH)-mono resistance), HIV status, and previous treatment history for tuberculosis, including new cases, relapse, loss to follow-up, and treatment failure, were used as independent variables.

### Statistical analysis

The researchers entered the data into the standard form and exported it to Excel for convenience to import and analyze using SPSS version 26. They performed descriptive summary statistics to evaluate the completeness of the data; assessed the proportional hazard assumption using graphical methods and Schoenfeld residual tests; ran Akaike information criterion to select model best fit the data; conducted bi-variable and multi-variable analysis using Cox regression model; finally, reported the strength of the association using the adjusted Hazard Ratio (aHR), along with its corresponding 95% Confidence Interval (CI) and a significance level of 5%.

### Ethical principles

The team sought written Ethical approval for the study from Hawassa University’s Institutional Review Board (IRB) and a support letter from the Sidama Regional Public Health Institute. They submitted the ethical clearance records to the Yirgalem Hospital. As the study was retrospective, we could not find the patients to consent in person. Consequently, the researchers asked the head of the Multidrug-resistant tuberculosis treatment initiation center to consent on behalf of the patients to access and extract data from the medical records. In addition, the study team discussed with the treatment initiation center staff to ensure transparency on study purposes, methods, and benefits of the study in improving the quality of the health services. The team de-identified the data collection form and kept it securely in locked cabinets; in addition, they kept the database password-protected to ensure the confidentiality of the patients. In general, we conducted the study per the 1964 Declaration of Helsinki.

### Participant agreement

Due to the retrospective nature of this follow-up study, it was not feasible to identify individual patients. However, we secured written informed consent from the director of the Yirgalem MDR-TB treatment-initiating center to proceed with the research.

## Results

A total of 420 patients with Multidrug-resistant tuberculosis enrolled over an 11-year follow-up period. Among the patients enrolled, 74 were excluded for not meeting the criteria, 55 for incomplete records, 18 for the age below fifteen, and 1 for an extra-pulmonary patient. Finally, the study analyzed 346 patients. Among the 346 participants, 225 (65%) were male and 121 (35%) were female. The mean and median ages of the participants were 30 and 27 years, respectively, with a standard deviation of ±11.78 (Table 3).

**Table 2:** The sample size calculation output for the study.

| Parameters | Kelsey | Fleiss | Fleiss with continuity correction |
| --- | --- | --- | --- |
| Sample Size - Exposed | 1888 | 1886 | 1977 |
| Sample Size-Non-exposed | 1888 | 1886 | 1977 |
| Total sample size: | 3776 | 3772 | 3954 |

**Table 3:** Baseline features of the patients at the time of initiation of second-line anti-TB treatment in the Sidama region, Ethiopia, from January 1, 2013 to June 30, 2024 (n =346).

| Variables | Frequency | Percent |
| --- | --- | --- |
| Age category | Frequency | Percent |
| 15-24 | 109 | 31.5 |
| 25-34 | 135 | 39.0 |
| 35-44 | 51 | 14.7 |
| 45-54 | 32 | 9.2 |
| 55-64 | 15 | 4.3 |
| >65 | 4 | 1.2 |
| Total | 346 | 100 |
| HIV Status |  |  |
| HIV-positive | 31 | 8.96 |
| HIV-negative | 313 | 90.5 |
| Unknown | 2 | 0.6 |
| Total | 346 | 100 |
| Resistance type |  |  |
| RR | 329 | 95.1 |
| MDR | 15 | 4.3 |
| INH resistance | 2 | .6 |
| Total | 346 | 100.0 |
| Patient registration group |  |  |
| New | 65 | 18.8 |
| Relapse | 160 | 46.2 |
| Loss to follow-up | 51 | 14.7 |
| Treatment failure | 70 | 20.2 |
| Total | 346 | 100.0 |
| Treatment regimen |  |  |
| Short term treatment | 69 | 19.9 |
| Long term treatment | 266 | 76.9 |
| Standardized | 3 | .9 |
| Individualized | 2 | .6 |
| BPaLM | 6 | 1.7 |

### Time-to-sputum culture conversion

Among the 346 study participants, 302 (87.3%) achieved sputum-culture conversion in a median time of 76 days (95% CI: 71-79, p < 0.05); on top of that, 36 patients (10.4%) displayed a delayed conversion, with a median conversion time of 141 days (95% CI: 132-151, p < 0.05). On the other hand, 8 (2.3%) patients did not show culture conversion by the end of the treatment (95% CI: 0.023, 0.01-0.04, p < 0.05). The follow-up period for participants totaled 29,001 person-days. The cumulative probabilities of survival or sputum culture non-conversion at the end of the second, third, fourth, and sixth months were 0.90, 0.55, 0.22, and 0.15, respectively. (Figure 1).

**Figure 1:**
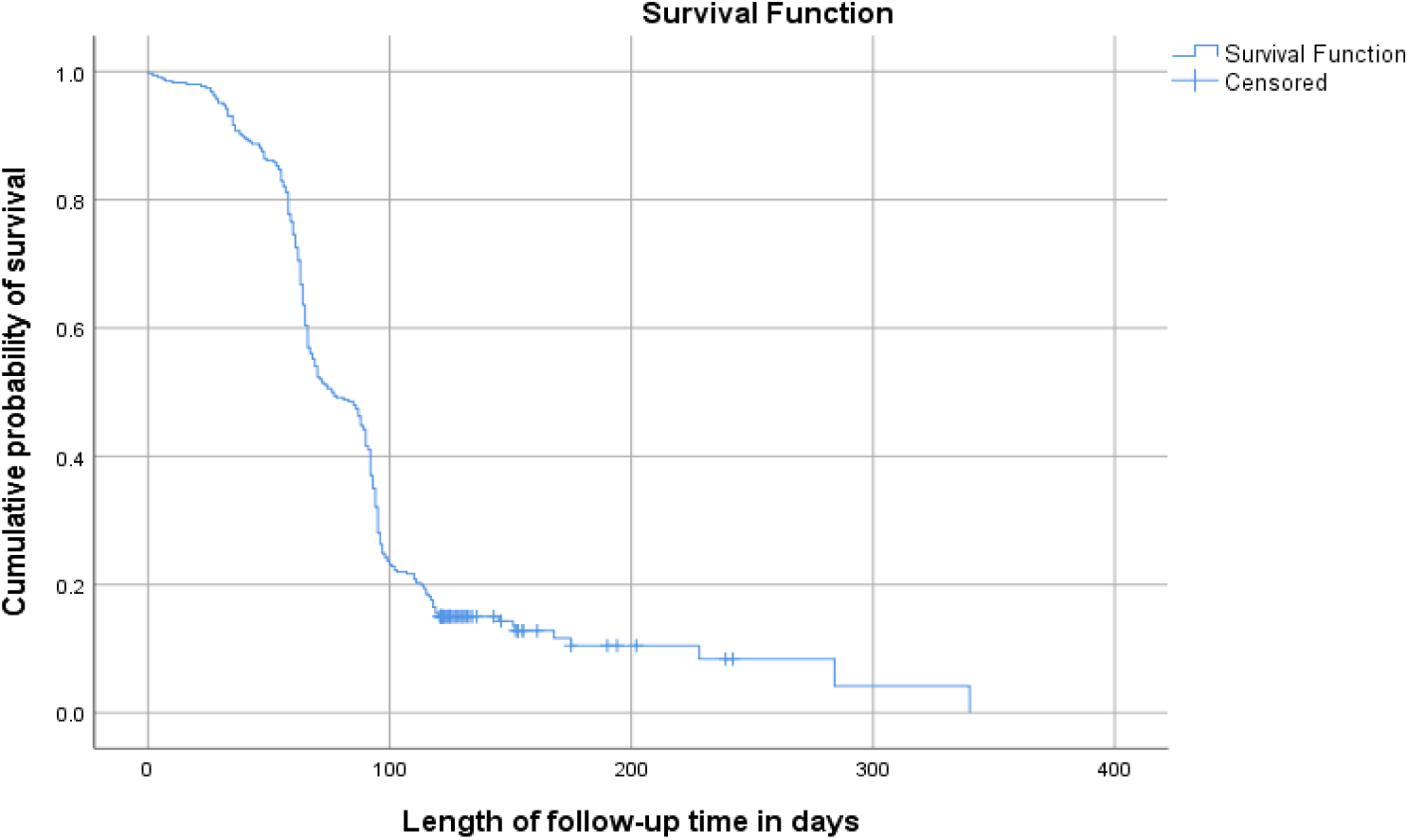
Kaplan-Meyer graph of time to sputum culture conversion among Multidrug-resistant tuberculosis patients in the Sidama region, from January 1, 2013 to June 30, 2024.

### Factors associated with time-to-sputum culture conversion

With 95% CI, Compared with males, females had a lesser likelihood of early time-to-sputum culture conversion, with an aHR of 0.035 (0.004-0.28), P = 0.001.

In terms of HIV status, people living with HIV had an earlier sputum culture conversion compared with HIV-negative people, with an aHR of 15.6 (4.5-54), P = 0.001.

People who registered with a drug-resistance type of MDR-TB were associated with early time-to-sputum culture conversion with aHR 4.7 (1-21), P = 0.041 compared with the patients with a resistance type of RR-TB, while a treatment regimen of long-term MDR-TB treatment was significantly associated with an aHR of 10.1 (2.2-45), P = 0.003 compared with the short-term treatment.

Nevertheless, different age categories and patient registration groups, like new, relapse, loss to follow-up, and treatment failure were not associated with early time-to-sputum culture conversion.

Factors that associated with time-to-sputum culture conversion were given bellow (Table 2).

### Treatment outcomes

Among the 346 study participants, 32 patients (9.2%) were not evaluated for their treatment outcomes. Of these, 26 patients had not completed their treatment course, while 6 patients had shifted to other facilities.

The overall treatment success rate was 234 (67.6%). The cure and treatment completion rates were 177 (51.2%) and 57 (16.5%), respectively. In contrast, the rate of unfavorable treatment outcomes was 80 (23%). Among these, 23 (6.6%) died, 5 (1.4%) experienced treatment failure, and 52 (15%) cases were lost to follow-up (Figure 3).

**Figure 2:**
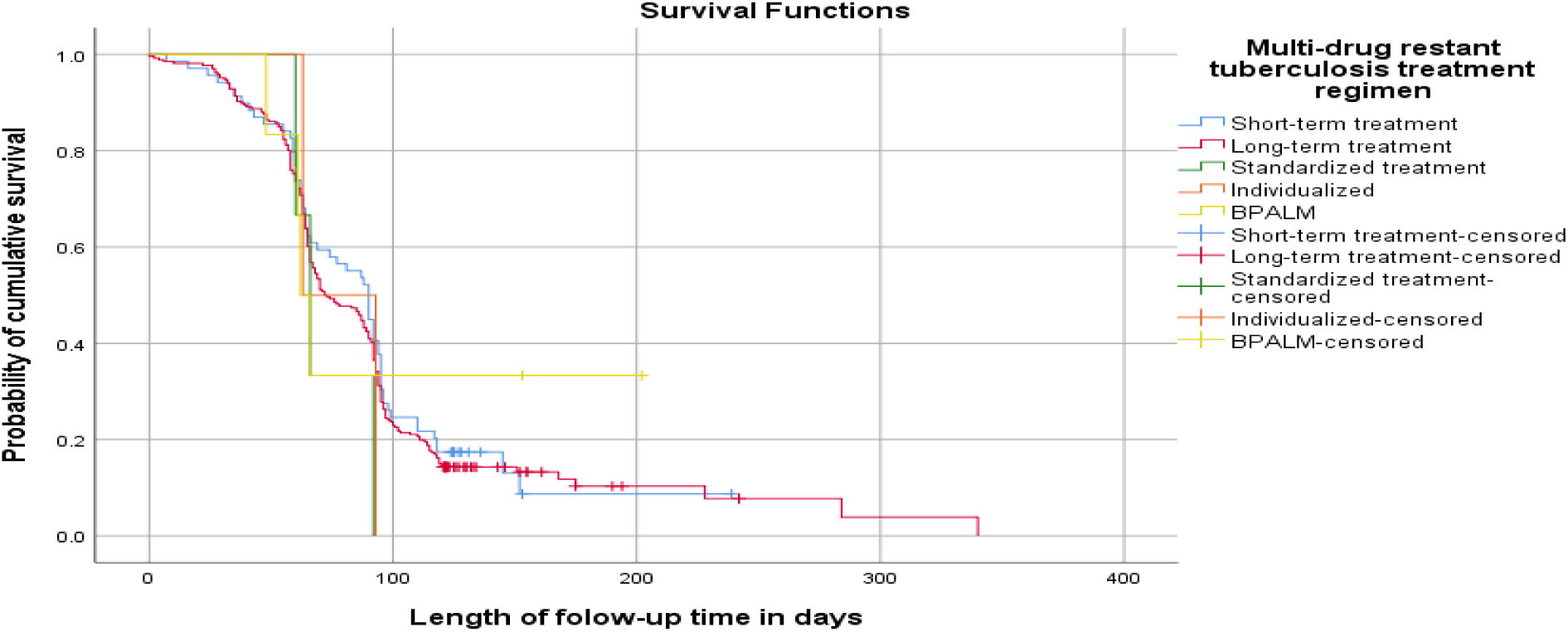
Kaplan-Meyer graph of time-to-culture conversion among multidrug-resistant tuberculosis patients per treatment regimen in the Sidama region, from January 1, 2013 to June 30, 2024.

**Figure 3:**
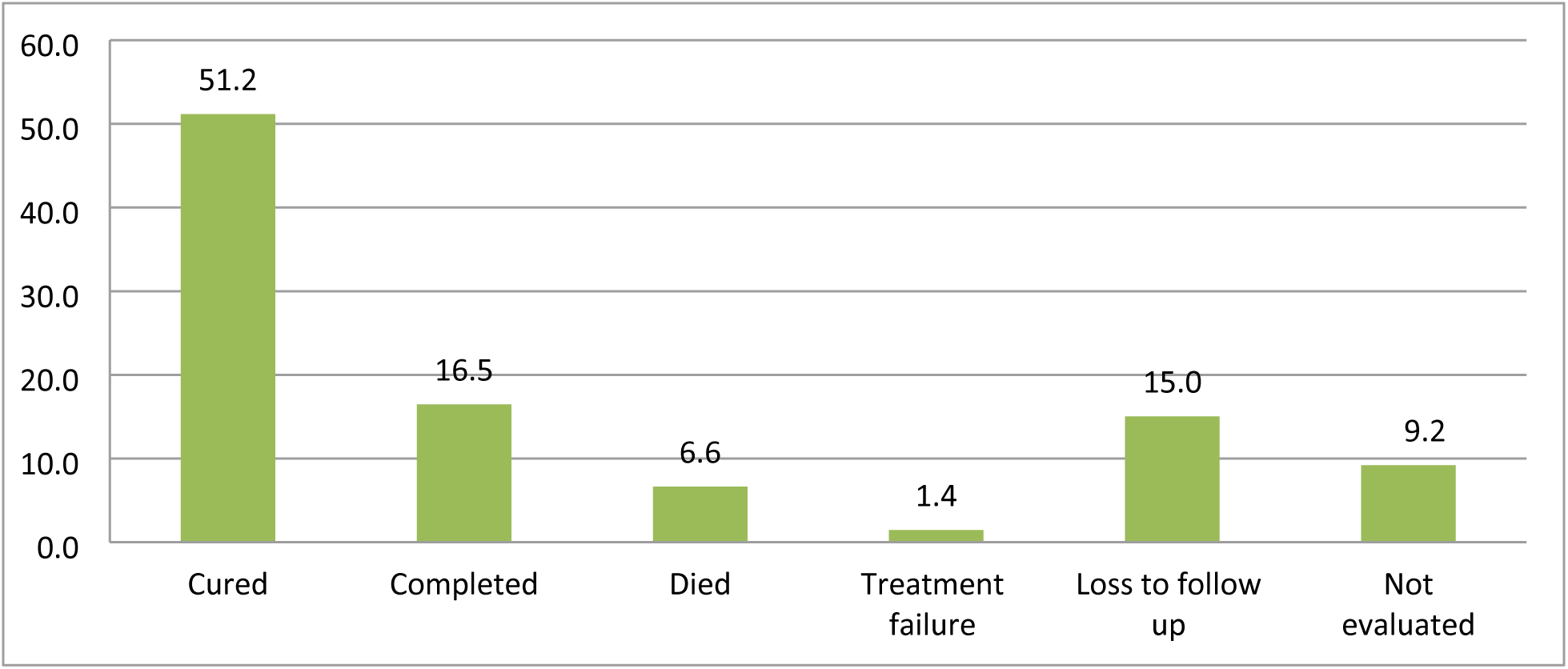
Treatment outcomes in multidrug-resistant tuberculosis patients in the Sidama region, from January 1, 2013, to June 30, 2024

### Factors associated with treatment outcomes

With 95% CI, compared with HIV-negative people, people living with HIV were associated with unfavorable treatment outcomes with aHR 12 (1-160), P = 0.047.

In terms of patient registration groups, patients who had previous treatment failure history were associated with unfavorable treatment outcomes compared to new patients, with an adjusted hazard ratio (aHR) of 6 (1-27), P = 0.0022.

On the other hand, in this study, different sex, age categories, drug resistance types, treatment regimens, and culture reversion were not significantly associated with unfavorable treatment outcomes.

### Sputum culture reversion

Among the 302 study participants who achieved initial sputum culture conversion, 5 (1.66%) participants had experienced culture reversion during their treatment course. In this study, culture reversion was not significantly associated with the overall unfavorable treatment outcome. However, all of the 5 participants who experienced culture reversion after initial culture conversion had also faced treatment failure (Table 6).

**Table 4:** Factors associated with the time-to-culture conversion among multidrug-resistant tuberculosis patients in the Sidama region from January 1, 2013 to June 32, 2024 (n= 346).

| Variables (n=346) | N (%) | Culture conversion N (%) |  | CHR | p-value | AHR | p-value |
| --- | --- | --- | --- | --- | --- | --- | --- |
| Sex | Total N= 346 | Event N= 302 | Censored |  |  |  |  |
| Male | 225 (65) | 195 (65) | 30 (68) | Reference |  |  |  |
| Female | 121(35) | 107 (35) | 14 (32) | 0.045 | .003 | 0.035 (0.004-0.28) | 0.001 |
| Age |  |  |  |  |  |  |  |
| 15 -24 | 109 (32) | 89 (29) | 20 (45.5) | Reference |  |  |  |
| 25-34 | 135 (39) | 124 (41) | 11 (25) | 1.4 (0.7-3) | .388 | 1.5 (0.68-3) | 0.34 |
| 35-44 | 51 (15) | 47 (16) | 4 (9) | 1.4 (0.7-3) | .390 | 1.2 (0.5-.2.7) | 0.63 |
| 45-54 | 32 (9.2) | 28(9.3) | 4 (9) | 1.2 (0.5-2.6) | .669 | 1.13 (0.5-2.6) | 0.78 |
| 55-64 | 15 (4.3) | 11(4) | 4 (9) | .85 (0.3-2) | .727 | 1 (0.4-2.6) | 0.99 |
| >65 | 4 (1.2) | 3(1) | 1 (2.3) | 1.0 (0.3-4) | .924 | 1.22 (0.3-4.8) | 0.77 |
| HIV status |  |  |  |  |  |  |  |
| HIV-negative | 313 (90) | 274 | 39 | Reference |  |  |  |
| HIV-positive | 31 (9) | 26 | 5 | 0.2 (0.1-0.5) | .001 | 15.6 (4.5-54) | 0.0 |
| Unknown | 2 (1) | 2 | 0 | 0.4(0.1-2.8) | 0.4 | 0.78 (0.02-35) | 0.9 |
| Drug-resistance type |  |  |  |  |  |  |  |
| RR-TB | 329 (95) | 286 | 43 | Reference |  |  |  |
| MDR-TB | 15 (4.3) | 14 | 1 | 4 (1.7-4) | .002 | 4.7 (1-21) | 0.041 |
| INH mono resistance | 2 (0.6) | 2 | 0 | 2.4 (0.5-10) | 0.3 | 1.7 (0.13-23) | 0.69 |
| Registration group |  |  |  |  |  |  |  |
| Treatment failure | 70 (20) | 61 | 9 | Reference |  |  |  |
| New | 160 (46) | 141 | 19 | 0.5 (0.2-1) | 0.36 | 0.47 (0.23-0.98) | 0.37 |
| Relapse | 65 (19) | 52 | 13 | 0.71 (0.4-1.4) | 0.35 | 0.7 (36-1.4) | 0.33 |
| Loss to follow-up | 51 (15) | 48 | 3 | 1 (0.5-2) | .877 | 0.99 (0.48-2.1) | 0.98 |
| MDR-TB treatment |  |  |  |  |  |  |  |
| Short-term | 69 (20) | 59 | 10 | Reference |  |  |  |
| Long-term | 266 (77) | 234 | 32 | 1.7 (0.79-3.3) | 0.15 | 10.1 (2.2-45) | 0.003 |
| Standardized | 3 (1) | 3 | 0 | 2.7 (07-10) | 0.1 | 4.4 (2-9) | 0. 06 |
| Individualized | 2 (0.6) | 2 | 0 | 2.3 (0.72-11) | 0.3 | 6.9 (1.4-205) | 0.27 |
| BPALM | 6 (1.7) | 4 | 2 | 1.2 (0.35-3.9) | 0.8 | 7.7 (0.84-70) | 0.07 |

**Table 5:** Factors Associated with Unfavorable Treatment Outcomes among the study participants (n = 346). N.B.: 32 patients were not evaluated for their treatment outcomes; therefore, treatment success was assessed in 314 patients.

| Variables (n=346) | N (%) | Treatment outcome N (%) |  | CHR | p-value | AHR | p-value |
| --- | --- | --- | --- | --- | --- | --- | --- |
| Sex | Total N= 314 | Unfavorable N=80 | Favorable N = 234 |  |  |  |  |
| Male | 204(65) | 60(19 ) | 144 (46) | Reference |  |  |  |
| Female | 110(35) | 20 (6) | 90(29) | 0.18 (0.1-1) | 0.67 |  |  |
| Age |  |  |  |  |  |  |  |
| 15 -24 | 97(31) | 26(8) | 71 ( 23 ) | Reference |  |  |  |
| 25-34 | 122(39) | 33(11) | 89 (28) | .95(0.2-5) | 0.94 | .98 (0.2-5) | 0.98 |
| 35-44 | 48(15) | 13(4) | 35 (11) | 1.1 (0.2-6) | 0.93 | 1.5 (0.3-9) | 0.64 |
| 45-54 | 29 (9) | 4(1) | 25 (8) | 2.3 (0.4 -4) | 0.37 | 2.3(0.3-15) | 0.39 |
| 55-64 | 14 (5) | 3(1) | 11 (4) | 1.3 (0.2-10) | 0.78 | 1.8 (0.2-16) | 0.61 |
| >65 | 4(1) | 1(0.3) | 3 (0.95) | 1.0 (0.1-10) | 0.95 | 1.3 (0.1-22) | 0.84 |
| HIV status |  |  |  |  |  |  |  |
| HIV-negative | 282(90) | 69 ((22) | 213 (68) | Reference |  |  |  |
| HIV-positive | 31(9.8) | 10(3.2) | 21 (6.7) | 15.9(1.7-148) | 0.015 | 12 (1-160) | 0.047 |
| Unknown | 1(0.3) | 1(0.3) | 0 |  |  |  |  |
| Drug-resistance type |  |  |  |  |  |  |  |
| RR-TB | 300 (95.5) | 78 (25) | 222 (71) | Reference |  |  |  |
| MDR-TB | 13 (4.1) | 2 (0.6) | 11 (3.5) | .176 (0.02-2) | .158 | 0 (00) | 0.99 |
| INH mono-resistance | 1 (0.3) | 0 | 1 (0.3) | .198 (0.00) | 1.000 | 10 (00) | 1 |
| Registration group |  |  |  |  |  |  |  |
| New | 64 (20) | 16 (5) | 48 (15) | Reference |  |  |  |
| Relapse | 145 (46) | 40 (13) | 105 (33) | 3.3 (0.9-13) | .079 | 3 (0.78-13) | 0.12 |
| Loss to follow-up | 38 (12) | 11 (3.5) | 27 (8.5) | 3 (0.7-13) | .128 | 3 (0.6-15) | 0.16 |
| Treatment failure | 67 (21) | 13 (4) | 54 (17) | 6 (1.4-25) | .015 | 6 (1-27) | 0.022 |
| MDR-TB treatment |  |  |  |  |  |  |  |
| Short-term | 64 (20) | 13 (4) | 51 (16) | Reference |  |  |  |
| Long-term | 242 (77) | 61 (19) | 181 (58) | .16 (0- 2) | .153 | 0.6 (0.05-7) | 0.7 |
| Standardized | 3 (1) | 2 (0.6) | 1 (0.3) | .03 ( 0-0.9) | .043 | 0.11 (0-3.6) | 0.22 |
| Individualized | 1 (0.3) | 0 | 1 (0.3) | 0 (0.0) | 1.0 | 00 (0.0) | 1 |
| BPALM | 4 (1) | 4 (1) | 0.0 | 0 (0.0) | 0.99 | 00 (0.0) | 0.99 |
| Culture reversion |  |  |  |  |  |  |  |
| Culture remained negative | 309 (98.4) | 75 (23.9) | 234 (74.5) | Reference |  |  |  |
| Culture reversed | 5 (1.6) | 5 (1.6) | 0.0 (0.0) | 0.0 (0.0) | 0.99 | 0.0 (0.0) | 0.99 |

**Table 6:** Culture reversion versus Treatment Outcomes among Multidrug-resistant tuberculosis Patients in the Sidama Region from January 1, 2013, to June 30, 2024 (n = 346).

| Treatment outcomes | Culture reversion |  | Total |
| --- | --- | --- | --- |
|  | Yes | No |  |
| Cured | 0 | 177 | 177 |
| Completed | 0 | 57 | 57 |
| Died | 0 | 23 | 23 |
| Treatment failure | 5 | 0 | 5 |
| Loss to follow-up | 0 | 52 | 52 |
| Not evaluated | 0 | 32 | 32 |
| Total | 5 | 341 | 346 |

## Discussions

Some of the previous studies have reported the initial median time-to-sputum culture conversion among multidrug-resistant tuberculosis in less than or equal to two months.^12-17^

On the other hand, the current study revealed that the median initial time to sputum culture conversion is 76 days, which is in agreement with other study findings that reported the initial median time-to-sputum culture conversion was approximately between two to three months.^4-6,18--25^

Consequently, the current study finding complies with the World Health Organization’s recommendation that the initial median time-to-sputum culture conversion to multidrug-resistant tuberculosis would be less than four months for a favorable treatment outcome prognosis.^6^

In the current study, the cumulative probabilities of survival or sputum culture non-conversion at the end of the second, third, fourth, and sixth months were 0.90, 0.55, 0.22, and 0.15, respectively. However, in the previous study, the cumulative probabilities of survival in the first, second, and fourth months were 0.89, 0.56, and 0.19, respectively.^22^

As a result, in the current study, the probabilities of sputum culture non-conversion were higher in the second and fourth months compared with the previous study findings. The finding signified that patients remained infectious for a longer time compared to the previous study.^22^

In this study, females were less likely than males to experience early culture conversion. However, a previous study found no significant association between sex categories and sputum culture conversion.

There was no significant association between initial time-to-sputum culture conversion with any age groups in the current study; however, the previous study reported that delayed sputum culture conversion was associated with old age.^27^

The current study, in agreement with many previous studies, revealed that people living with HIV had an earlier sputum culture conversion compared with HIV-negative people..^15, 19, 24, and 28^

The potential justification for this might be, in people living with HIV, due to immunity suppression, the patients fail to form cavitation that might have led to a false negative sputum culture result. In contrast, people living with HIV had worse treatment outcomes compared with HIV-negative people.

On the other hand, some other previous studies reported that there was no significant association between time-to-sputum culture conversion and HIV status.^11, 26^

In the current study, patients registered with a drug-resistant type of MDR-TB experienced a shorter time-to-sputum culture conversion compared to those with RR-TB. However, the previous study reported the absence of an association with any drug resistance type.^24^

The current study disclosed that time-to-sputum culture conversion was not associated with any patient registration group. In contrast, the previous study had reported that the patient registration group being new to second-line anti-TB treatment was associated with earlier time-to-sputum culture conversion.^4, 6^

In the current study, compared with short-term MDR-TB treatment, the long-term treatment was associated with an earlier time-to-sputum culture conversion. In contrast, a previous study reported that drug regimens containing kanamycin were associated with a prolonged time.^18^

Out of the 346 participants in the study, we did not evaluate 32 individuals: 26 did not complete the treatment, and 6 transferred out of the study area.

The overall treatment success rate was 234 (67.6%). The cure and treatment completion rates were 177 (51.2%) and 57 (16.5%), respectively. In contrast, the rate of unfavorable treatment outcomes was 80 (23%). Among these, 23 (6.6%) died, 5 (1.4%) experienced treatment failure, and 52 (15%) cases were lost to follow-up.

Compared to the previous study that exposed the overall treatment success, cure, and treatment completion rates of 154/189 (81%), 131/189 (54%), and 23/189 (10%), respectively. The treatment success rate was lower in the current study.^30^

In comparison to previous studies that reported unsuccessful treatment outcomes, of 31/189 (13%), and 4/189 (2%), died, and treatment failure respectively, the current study demonstrated significantly better and lower findings.^31^

However, compared to another earlier study that reported the unfavorable rate of treatment outcomes, 3 (4.4%) and 6 (8.8%) died, and lost-to-follow-up respectively, the current rates were higher.^32^

Nonetheless, the rate of loss to follow-up was worse in the current study than in the previous study reports.^32,^

On the other hand, out of the 302 patients who achieved early culture conversion, 5 (1.66%) experienced culture reversion. However, the culture reversion rate was lower compared to the previous study, which had reported that 54/1286 (4.2 %).^26^

Sputum culture reversion after conversion serves as an indicator of TB treatment failure. A preceding study found that among patients who had culture reversion, a significant majority (92.6%; n = 50) did not achieve successful end-of-treatment outcomes.^26^

In this regard, all patients who experienced culture reversion (5/5, 100%) also faced treatment failure.

## Conclusions

A vast majority of patients (87.3%) achieved sputum culture conversion, highlighting the effectiveness of the treatment for MDR-TB within the study population.

Consequently, the median time to early sputum-culture conversion for the patients was 76 days, while the remaining subsets of patients (10.4%) experienced a delayed conversion, with a median time of 141 days, and (2.3%) of the patients did not show culture conversion.

The findings from the current study indicate that patients experienced higher probabilities of sputum culture non-conversion in the second and fourth months compared to the previous findings. The study finding suggests that patients may have remained infectious for a longer duration in the current study, which could have implications for treatment strategies and public health measures.

In particular, female patients exhibited a lesser likelihood of early sputum-culture conversion compared to males, suggesting potential gender-related factors influencing treatment response.

On the other hand, Individuals living with HIV had notably earlier sputum-culture conversion compared with those who were HIV-negative.

However, in the same study, the MDR-TB treatment outcome analysis has shown that people living with HIV were associated with an unfavorable treatment outcome, leaving after a further study question. In addition, patients with a history of treatment failure faced unfavorable outcomes more frequently than new patients, pointing to serious follow-up of the MDR-TB during the treatment process.

Moreover, patients with the drug-resistance type of MDR-TB experienced earlier conversion than those with rifampicin-resistant tuberculosis (RR-TB), suggesting close monitoring of the treatment per the patient’s resistance type.

Furthermore, patients undergoing long-term MDR-TB treatment regimens demonstrated a greater likelihood of early conversion compared to those on short-term regimens, emphasizing the importance of treatment duration in patient outcomes.

## Recommendations

Address the lower likelihood of early sputum-culture conversion in female patients, considering potential social, psychological, or biological factors.

The current study highlights a concerning trend in which patients show higher probabilities of sputum culture non-conversion. The finding could indicate a need for public health interventions to mitigate the risk of transmission.

Tailor monitoring for all MDR-TB patients to minimize treatment failure and to ensure favorable treatment outcomes.

Emphasize the importance of treatment duration based on individual patient needs.

Long-term regimens have a higher likelihood of early culture conversion, signifying the appropriateness.

## Limitations of the study

As this is a retrospective follow-up study, we encountered challenges related to incomplete information during data collection. We were unable to obtain comprehensive patient details, including marital status, education level, religion, baseline alcohol consumption, smoking habits, khat chewing, liver function tests, and renal function tests. Additionally, some patients did not have regular monthly sputum culture examinations.

## Supporting Information

S1 Data collection form

## Data Availability

All relevant data are within the manuscript and its Supporting Information files.

## Acknowledgements

I thank the Hawassa University Public Health, Academic, and Service Directorate Office for the guidance and advice I have gotten from the office.

I would like to thank the leadership and staff of the Sidama Regional State Health Bureau, Sidama Public Health Institute, and Yirgalem General Hospital for their support.

My heartfelt goes to Dr. Kebede Tefera for his insightful comment and invaluable advice during the study period, to Mr. Adato Adela (Regional Laboratory Director), Mr. Erdachew Ambaye (Quality Assurance Laboratory Section Supervisor), Dagim Abebayehu, Abinet Alemayehu, and Dinknesh Argaw, for their unreserved assistance during the research data collection.

## Authors’ contributions

Conceptualization: Wolde Abreham Geda

Data curation: Wolde Abreham Geda, Kebede Tefera Betru,,Solomon Daniel

Formal analysis: Wolde Abreham Geda, Kebede Tefera Betru, Tarekegn Solomon

Funding acquisition : Wolde Abreham Geda

Investigation: Wolde Abreham Geda, Kebede Tefera Betru, Tarekegn Solomon, Solomon Daniel

Methodology: Wolde Abreham Geda, Kebede Tefera Betru, Tarekegn Solomon, Solomon Daniel

Resources: Wolde Abreham Geda

Software : Wolde Abreham Geda

Supervision: Wolde Abreham Geda Kebede Tefera Betru,Tarekegn Solomon

Validation : Wolde Abreham Geda, Kebede Tefera Betru, Tarekegn Solomon, Solomon Daniel

Visualization : Wolde Abreham Geda, Kebede Tefera Betru, Tarekegn Solomon

Writing – original draft: Wolde Abreham Wolde Abreham Geda, Kebede Tefera Betru

Writing – review & editing: Wolde Abreham Geda, Kebede Tefera Betru, Tarekegn Solomon, Solomon Daniel

## References

1. Global tuberculosis report 2024. Geneva: World Health Organization; 2024. Licence: CC BY-NC-SA 3.0 IGO.

2. World health statistics 2024: monitoring health for the SDGs, Sustainable Development Goals. Geneva: World Health Organization; 2024. Licence: CC BY-NC-SA 3.0 IGO.

3. Global tuberculosis report. Geneva: World Health Organization; 2023. License: CC BY-NC-SA 3.0 IGO.

4. Yihunie Akalu T, Muchie KF, Alemu Gelaye K. Time to sputum culture conversion and its determinants among Multidrug-resistant Tuberculosis patients at public hospitals of the Amhara Regional State: A multicenter retrospective follow up study. PLoS ONE 2018. 13(6):e0199320. 10.1371/journal.pone.0199320.

5. Li, Qingchun et al. Time to sputum culture conversion and its predictors among patients with multidrug-resistant tuberculosis in Hangzhou, China: A retrospective cohort study. MedicineBaltimore). 2020Dec11; 99(50):e23649. doi: 10.1097/MD.0000000000023649. PMID: 33327347; PMCID: PMC7738096.

6. Shibabaw A, Gelaw B, Wang S-H, Tessema B. Time to sputum smear and culture conversions in multidrug resistant tuberculosis at University of Gondar Hospital, Northwest Ethiopia. PLoS ONE 13(6): e0198080.10.1371/journal.pone.0198080.

7. Liu Q, Lu P, Martinez L, Yang H, Lu W, Ding X, Zhu L. 2018. Factors affecting time to sputum culture conversion and treatment outcome of patients with multidrug-resistant tuberculosis in China. BMC Infect Dis. 2018 Mar 6;18(1):114. doi: 10.1186/s12879-018-3021-0. PMID: 29510666; PMCID: PMC5840772.

8. Kelsey JL. Methods in Observational Epidemiology. Monographs in Epidemiology,1996.

9. Fleiss JL, Levin B, Paik MC. Statistical methods for rates and proportions. John Wiley & Sons; 2013 Jun 12.

10. Girum T, Muktar E, Lentiro K, Wondiye H, Shewangizaw M. Epidemiology of multidrug-resistant tuberculosis (MDR-TB) in Ethiopia: a systematic review and meta-analysis of the prevalence, determinants and treatment outcome. Tropical diseases, travel medicine and vaccines. 2018 Dec;4:1–2.

11. Dean AG, Sullivan KM, Soe MM. OpenEpi: Open Source Epidemiologic Statistics for Public Health, Version. www.OpenEpi.com, updated 2013/04/06, accessed 2024/02/10.

12. Mpagama, S.G. et al. Diagnosis and interim treatment outcomes from the first cohort of multidrug-resistant tuberculosis patients in Tanzania. PloS one, 2013 8(5), p.e62034.

13. Putri FA.et al. Body mass index predictive of sputum culture conversion among MDR-TB patients in Indonesia. The International journal of tuberculosis and lung disease. 2014 May 1;18 (5):564–70.

14. Meshesha MD. Predictors of sputum culture conversion time among MDR/RR TB patients on treatment in a low-income setting. PLoS One. 2022 Nov 14; 17(11):e0277642.doi: 10.1371/journal.pone.0277642. PMID: 36374857; PMCID: PMC9662721.

15 Lv, Lingshuang, et al. Sputum bacteriology conversion and treatment out-come of patients with multidrug-resistant tuberculosis .Infection and drug resistance.2018 Jan 1:147–54.

16. Bade AB, Mega TA, Negera GZ. Malnutrition is Associated with Delayed Sputum Culture Conversion Among Pati ents Treated for MDR-TB. Infect Drug Resist. 2021 Apr 28;14:1659–1667. doi: 10.2147/IDR.S293461. PMID: 33953577; PMCID: PMC8089472.

17. Ncha R. et al. Predictors of time to sputum culture conversion in Multidrug-resistant tuberculosis and extensively drug-resistant tuberculosis in patients at Tshepong-Klerksdorp Hospital. S Afr J Infect Dis. 2019 Aug 26;34(1):111. doi: 10.4102/sajid.v34i1.111. PMID: 34485452; PMCID: PMC8377786.

18. Megerso A. et al. A retrospective comparative study on median time to sputum culture conversion in multidrug-resistant pulmonary tuberculosis patients in pastoral and non-pastoral settings in southeast oromia, Ethiopia. Infection and Drug Resistance. 2021 Dec 14:5325–33.

19. Brust JCM.et al. Chest Radiograph Findings and Time to Culture Conversion in Patients with Multidrug-Resistant Tuberculosis and HIV in Tugela Ferry, South Africa. PLoS ONE. 2013 8(9): e73975. doi:10.1371/journal.pone.0073975.

20. Bastard M.et al. What is the best culture conversion prognostic marker for patients treated for multidrug-resistant tuberculosis? The International Journal of Tuberculosis and Lung Disease. 2019 Oct 1; 23 (10):1060–7.

21. Muvunyi CM. et al. Highly successful treatment outcome of multidrug-resistant and genetic diversity of multidrug-resistant Mycobacterium tuberculosis strains in Rwanda. Tropical Medicine & International Health. 2019 Jul;24(7):879–87.

22. Weldemhret Let al. Time to Sputum Culture Conversion and Its Predictors Among Multidrug Resistant Tuberculosis Patients in Tigray, Northern Ethiopia: Retrospective Cohort Study. Infection and Drug Resistance. 2023 Dec 31:3671–81.

23. Magee MJ., et al. Diabetes Mellitus, Smoking Status, and Rate of Sputum Culture Conversion in Patients with Multidrug-Resistant Tuberculosis: A Cohort Study from the Country of Georgia. PLoS ONE 2014 9(4): e94890. doi:10.1371/journal.pone.0094890.

24. Tekalegn Y. et al Predictors of Time to Sputum Culture Conversion Among Drug-Resistant Tuberculosis Patients in Oromia Region Hospitals, Ethiopia. Infect Drug Resist. 2020 Jul 27;13:2547–2556. doi: 10.2147/IDR.S250878. PMID: 32821129; PMCID: PMC7419643.

25. Velayutham B, Nair D, Kannan T, Padmapriyadarsini C, Sachdeva KS, et al. (2016) Factors associated with sputum culture conversion in multidrug resistant pulmonary tuberculosis. Int J Tuberc Lung Dis 20:1671–1676. PMID: 27931345.).

26. Kho S. et al. Sputum culture reversion in longer treatments with bedaquiline, delamanid, and repurposed drugs for drug-resistant tuberculosis. Nat Commun 15, 3927 (2024). 10.1038/s41467-024-48077-8.

27. Assemie MA. et al. Time to sputum culture conversion and its associated factors among multidrug-resistant tuberculosis patients in Eastern Africa: A systematic review and meta-analysis. Int J Infect Dis. 2020 Sep;98 :230–236. doi: 10.1016/j.ijid.2020.06.029. Epub 2020 Jun 12. PMID: 32535296.

28. Avaliani T. et al. Effectiveness and safety of fully oral modified shorter treatment regimen for multidrug-resistant tuberculosis in Georgia, 2019-2020. Monaldi Arch Chest Dis. 2021.Jan 14;91(1):10.4081/monaldi.2021.1679.doi:10.4081/monaldi.2021.1679. PMID: 33470088; PMCID: PMC9391985.

29. Results report 2024 [Internet]. https://www.theglobalfund.org/. Geneva : The Global Fund to Fight AIDS, Tuberculosis, and Malaria; 2024 Nov [cited 2025 Jan 1] p. 1–167. Available from: https://www.theglobalfund.org/en/about-the-global-fund/.

30. Alene, K.A. et al. Treatment outcomes in patients with multidrug-resistant tuberculosis in north-west Ethiopia. Tropical medicine & international health. 2017. 22(3), pp.351–362.

31. Martin MK. Et al. High rates of culture conversion and low loss to follow-up in MDR-TB patients managed at Regional Referral Hospitals in Uganda. BMC Infect Dis. 2021 Oct 12;21(1):1060. doi: 10.1186/s12879-021-06743-y. PMID: 34641816; PMCID: PMC8507334.

32. Kuchukhidze G et al. Risk factors associated with loss to follow-up among multidrug-resistant tuberculosis patients in Georgia. Public Health Action. 2014 Oct 21;4 (Suppl 2):S41-6. doi: 10.5588/pha.14.0048. PMID: 26393097; PMCID: PMC4547510.

